# Hydroxychloroquine is associated with slower viral clearance in clinical COVID-19 patients with mild to moderate disease: A retrospective study

**DOI:** 10.1101/2020.04.27.20082180

**Authors:** Jihad Mallat, Fadi Hamed, Maher Balkis, Mohamed A. Mohamed, Mohamad Mooty, Asim Malik, Ahmad Nusair, Maria-Fernanda Bonilla

## Abstract

**Background:** There is conflicting data regarding the use of hydroxychloroquine (HCQ) in COVID-19 hospitalized patients

**Objective:** To assess the efficacy of HCQ in increasing SARS-CoV-2 viral clearance

**Design:** Retrospective observational study

**Setting:** Cleveland Clinic Abu Dhabi

**Participants:** Hospitalized adult patients with confirmed SARS-CoV-2 infection

**Intervention:** None

**Measurements:** The primary outcome was the time from a confirmed positive nasopharyngeal swab to turn negative. A negative nasopharyngeal swab conversion was defined as a confirmed SARS-CoV-2 case followed by two negative results using RT-PCR assay with samples obtained 24 hours apart

**Results:** 34 confirmed COVID-19 patients were included. Nineteen (55.9%) patients presented with symptoms, and 14 (41.2%) had pneumonia. Only 21 (61.8%) patients received HCQ. The time to SARS-CoV-2 negativity nasopharyngeal test was significantly longer in patients who received HCQ compared to those who did not receive HCQ (17 [13–21] vs. 10 [4–13] days, p=0.023). HCQ was independently associated with time to negativity test after adjustment for potential confounders (symptoms, pneumonia or oxygen therapy) in multivariable linear regression analysis. On day 14, 47.8% (14/23) patients tested negative in the HCQ group compared to 90.9% (10/11) patients who did not receive HCQ (p=0.016).

**Limitations:** Small sample size and retrospective design with a potential risk of selection bias

**Conclusion:** HCQ was associated with a slower viral clearance in COVID-19 patients with mild to moderate disease. Data from ongoing randomized clinical trials with HCQ should provide a definitive answer regarding the efficacy and safety of this treatment.

## Introduction

Since December 2019, a novel coronavirus SARS-CoV-2 emerged in Wuhan city and rapidly spread throughout China [1]. Since then, the virus has extended around the world, crossing the Middle East and North Africa region, to Europe and then currently to North America, which has become the epicenter of the pandemic. As of April 19, 2020, a total of around 2,241,778 confirmed cases have been documented globally, with more than 152,551 deaths worldwide [2].

Therefore, the focus of therapeutic intervention has been to decrease the duration of viral shedding and thus limit the spread of the virus, and slow the progression of the disease. Besides of antiviral drugs, chloroquine and hydroxychloroquine (anti-malarial drugs) have been proposed as potential agents that could reduce the viral load and the transmission of the virus. Chloroquine analogs appear to block viral entry to cells by inhibiting the acidification of endosomes and glycosylation of host receptors [3–5]. Hydroxychloroquine (HCQ) has been demonstrated to be effective in inhibiting SARS-CoV-2 infection in-vitro studies [6,7].

Clinical studies have shown conflicting results. French studies suggested that HCQ, mainly when used with azithromycin, could reduce the viral load and improve the outcome of patients infected with SARS-CoV-2 [8,9]. Based on these results, HCQ has been prescribed off-label widely to improve the evolution of these patients. Even an international Task Force led by the American Thoracic Society suggests HCQ on a case-by-case basis for hospitalized patients with COVID-19 who have evidence of pneumonia [10]. However, the efficacy of HCQ in increasing viral clearance has been challenged in recent studies [11,12]. In addition, HCQ can induce QTc prolongation that could result in potentially severe cardiac dysrhythmia. Thus, this medication should not be used if it is not clinically proven as beneficial, in particular in COVID-19 patients with mild to moderate illness.

The aim of our study was to investigate the efficacy of early use of HCQ in increasing the viral clearance in confirmed hospitalized COVID-19 patients with mild to moderate disease.

## Materials and methods

### Patients

This was a retrospective observational study was performed at Cleveland Clinic Abu Dhabi. The institutional Ethics Committee of Cleveland Clinic Abu Dhabi approved the study, and a waiver of informed consent was obtained due to the nature of the retrospective chart review. All consecutive adult patients (≥18 years) admitted to our hospital between March 1 and 25, 2020, with confirmed SARS-CoV-2 infection were included. A confirmed case of SARS-CoV-2 was defined as a positive result of real-time reverse-transcriptase–polymerase-chain-reaction [RT-PCR] assay of a specimen collected on a nasopharyngeal swab according to the WHO guidance [13].

### Data collection

Deidentified data form the electronic medical record was collected. We obtained demographic data, information on clinical symptoms at presentation, and laboratory and radiological results during hospitalization. Imaging was reviewed by a specialized radiologist. C-reactive protein, ferritin level, white blood cells, neutrophil, and lymphocytes counts were also collected around day seven or at hospital discharge if the latter occurred before. Severe pneumonia was defined as the presence of pneumonia with the need for supplemental oxygen [13]. Time from hospital admission to onset pneumonia was also collected. The use of HCQ and the time from hospital admission to its administration were obtained. According to the hospital protocol, HCQ 400 mg was administered twice daily for 1 day, followed by 400 mg daily for 10 days.

A negative nasopharyngeal swab conversion was defined as a confirmed SARS-CoV-2 case followed by two negative results using RT-PCR assay with samples obtained 24 hours apart. Time to SARS-CoV-2 negativity test, which was our primary outcome, was calculated as the difference between the date of a second confirmed negative result and the date of the first confirmed positive test.

### Statistical analyses

No statistical sample size calculation was performed a priori, and the sample size was equal to the number of patients treated during the study period. Continuous variables are expressed as median and interquartile range (25–75%). Proportions are used as descriptive statistics for categorical variables. Comparisons of values between groups were performed using a Mann–Whitney *U* test. Pairwise comparisons between the different study periods were assessed using Wilcoxon’s test. Analyses of discrete data were performed using Fisher’s exact test. Simple linear regression analysis was performed to identify variables (HCQ treatment, symptoms, the presence of pneumonia, and oxygen therapy) that were associated with the time to negativity test. Multiple linear regression analysis was used to identify if HCQ was independently associated with the time to negativity test after adjusting for pneumonia or oxygen therapy, and symptoms. A bootstrap method with 1000 sampling with replacement was used to determine the 95% confidence intervals of regression coefficients parameters by the bias-corrected and accelerated bootstrap method [14].

Statistical analyses were performed using SPSS software version 20.0 (IBM corporation). p <0.05 was considered statistically significant. All reported p values were two-sided.

## Results

Thirty-four confirmed COVID-19 patients were enrolled. Among them, only 21 (61.8%) patients received HCQ. The median time from hospital admission to HCQ administration was 0 [0–2] days. The clinical characteristics of the patients are shown in Table 1. The median age was 37 years, and 73.5% were male. Comorbidities were found in 10 cases (29.4%) with essential hypertension being the most common. The median time from onset of symptoms to hospital admission was 4 days. The most common symptom on admission was cough (50%). Fever was present in only 23.5% of patients. Fourteen patients developed pneumonia. Among them, 6 (42.8%) patients required oxygen inhalation with a nasal cannula (2.5 [2.0–4.0] L/min). The median time from hospital admission to pneumonia was 1.0 [0.0–3.0] days (Table 1). No patients were admitted to intensive care unit, required high flow oxygen therapy, non-invasive or invasive mechanical ventilation, and all of them were discharged alive from the hospital.

**Table 1.** Comparisons of baseline characteristics and laboratory data between HCQ and non-HCQ groups

| Variables | All patients<br>(n=34) | HCQ<br>(n=23) | Non-HCQ<br>(n=11) | p-value |
| --- | --- | --- | --- | --- |
| Age (years) | 37 [31-48] | 33 [31-48] | 41 [30-55] | 0.64 |
| Male, n (%) | 25 (73.5) | 17 (73.9) | 8 (72.7) | 1.00 |
| Weight (kg) | 73 [65-83.] | 73 [64-82] | 75 [65-92] | 0.36 |
| BMI (kg/m<sup>2</sup>) | 24.4 [22.7-27.5] | 24.2 [21.2-26.6] | 24.9 [23.5-30.6] | 0.38 |
| Patients with comorbidities, n (%) | 10 (29.4) | 4 (17.4) | 6 (54.5) | **0.045** |
| Comorbidities distribution, n (%) |  |  |  |  |
| Asthma | 3 (8.8) | 2 (8.7) | 1 (9.1) |  |
| Diabetes | 2 (5.9) | 0 (0) | 2 (18.2) |  |
| Hypertension | 5 (14.7) | 0 (0) | 5 (45.5) |  |
| Malignancy | 3 (8.8) | 2 (8.7) | 1 (9.1) |  |
| Chronic heart failure | 1 (2.9) | 0 (0) | 1 (9.1) |  |
| Chronic kidney disease | 1 (2.9) | 0 (0) | 1 (9.1) |  |
| Immunosuppressive treatment | 1 (2.9) | 1 (4.3) | 0 (0) |  |
| Current tobacco smoker | 3 (8.8) | 1 (4.3) | 2 (18.2) |  |
| Non-steroidal anti-inflammatory treatment | 4 (11.8) | 3 (13) | 1 (9.1) | 1.00 |
| Patients with symptoms on admission, n (%) | 19 (55.9) | 12 (52.2) | 7 (63.6) | 0.71 |
| Duration of symptoms before admission (day) | 4.0 [2.0-7.0] | 4.0 [2.0-7.0] | 4.5 [1.2-6.5] | 0.65 |
| Symptoms distributions on admission, n (%) |  |  |  |  |
| Fever<sup>#</sup> | 8 (23.5) | 6 (26.1) | 2 (18.2) |  |
| Cough | 17 (50) | 10 (43.5) | 7 (63.6) |  |
| Sore throat | 6 (17.6) | 5 (21.7) | 1 (9.1) |  |
| Rhinorrhea | 5 (14.7) | 4 (17.4) | 1 (9.1) |  |
| Nasal congestion | 10 (29.4) | 7 (30.4) | 3 (27.3) |  |
| Shortness of breath | 6 (17.6) | 4 (17.4) | 2 (18.2) |  |
| Chest tightness | 5 (14.7) | 3 (13.0) | 2 (18.2) |  |
| Headache | 4 (11.8) | 1 (4.3) | 3 (27.3) |  |
| Fatigue | 14 (41.2) | 8 (34.8) | 6 (54.5) |  |
| Myalgia | 8 (23.5) | 6 (26.1) | 2 (18.2) |  |
| Diarrhea | 4 (11.8) | 1 (4.3) | 3 (27.3) |  |
| Dysgeusia and anosmia | 4/13 (30.8) | 4/11 (36.4) | 0/2 (0) |  |
| Highest temperature (°C) | 37.0 [36.9-37.4] | 37.0 [36.9-37.4] | 37.0 [36.37.2] | 0.77 |
| Highest heart rate (beats/min) | 87 [78-95] | 85 [74-94] | 93 [83-99] | 0.12 |
| Highest respiratory rate (breaths/min) | 19 [18-20] | 19 [18-20] | 18 [18-20] | 0.91 |
| Lowest arterial oxygen saturation on room air (%) | 98 [97-98] | 98 [97-98] | 97 [96-99] | 0.69 |
| Laboratory data on ICU admission |  |  |  |  |
| C-reactive protein (mg/L) | 3.7 [0.9-7.7] | 3.4 [0.7-7.7] | 4.3 [1.6-16.6] | 0.49 |
| Hemoglobin (g/L) | 146 [136-159] | 146 [138-159] | 148 [126-159] | 0.56 |
| Creatinine (μmol/L) | 77 [64-93] | 80 [67-98] | 70 [60-92] | 0.40 |
| Procalcitonin (ng/mL) | 0.04 [0.03-0.06] | 0.04 [0.02-0.05] | 0.06 [0.04-0.43] | **0.04** |
| Leucocytes count (/mm<sup>3</sup>) | 6045 [4590-7020] | 6170 [3820-6520] | 5920 [5160-7470] | 0.27 |
| Leucocytes ≥10,000/mm<sup>3</sup>, n (%) | 2 (5.9) | 0 (0) | 2 (18.2) | 1.00 |
| Leucocytes ≤ 4000/mm<sup>3</sup>, n (%) | 6 (17.6) | 6 (26.1) | 0 (0) | 0.14 |
| Lymphocytes count (/mm<sup>3</sup>) | 1670 [1167-1960] | 1650 [980-1950] | 1890 [1430-2230] | 0.42 |
| Lymphocytes ≤ 1500/mm<sup>3</sup>, n (%) | 13 (38.2) | 9 (39.1) | 4 (36.4) | 1.00 |
| Neutrophil/lymphocyte ratio | 2.07 [1.24-2.78] | 2.03 [1.42-2.78] | 2.50 [1.20-2.92] | 0.80 |
| Platelet count (/mm<sup>3</sup>) | 239 [177-272] | 236 [180-268] | 243 [167-284] | 0.74 |
| INR | 1.0 [1.0-1.1] | 1.1 [1.0-1.1] | 1.0 [1.0-1.1] | 0.91 |
| D-dimer (µg/mL) [normal reference:<0.05] | 0.32 [0.27-0.55] | 0.27 [0.27-0.40] | 0.54 [0.33-1.09] | **0.034** |
| Ferritin (µg/L) [reference range: 36-480] | 140 [49-322] | 165 [63-320] | 292 [33-1085] | 1.00 |
| Lactate dehydrogenase [reference range:135-225] | 209 [165-259] | 206 [162-238] | 265 [181-381] | 0.24 |
| Alanine aminotransferase (IU/L) | 26 [17-39] | 33 [15-40] | 21 [17-32] | 0.49 |
| Aspartate aminotransferase (IU/L) | 24 [20-31] | 23 [20-31] | 24 [20-43] | 0.83 |
| Bilirubin (µmol/L) [reference range: 5-21] | 8.9 [5.2-12.4] | 9.0 [5.0-12.4] | 7.6 [5.0-17.1] | 0.78 |
| Pneumonia, n (%) | 14 (41.2) | 11 (47.8) | 3 (27.3) | 0.29 |
| Time from admission to pneumonia (day) | 1.0 [0.0-3.0] | 0.0 [0.0-3.0] | 2.0 [0.0-2.0] | 1.00 |
| Oxygen inhalation, n (%) | 6 (17.6) | 4 (17.4) | 2 (18.2) | 1.00 |
| Hospital length of stay (day) | 14.0 [6.0-18.5] | 17.0 [6.0-20.0] | 9.0 [6.0-12.7] | 0.068 |

HCQ, hydroxychloroquine; BMI, body mass index

No significant differences were found in subject characteristics, symptoms rate, laboratory data, pneumonia rate, or oxygen therapy between HCQ and non-HCQ patients except for comorbidities rate and D-dimer levels, which were significantly higher in the non-HCQ group (Table 1). The hospital length of stay was longer in the HCQ group than in the non-HCQ group, but it did not reach statistical significance (p=0.068, Table 1). HCQ was well tolerated with no observed side effects.

### Factors associated with time to negativity test

The time to SARS-CoV-2 negativity test was significantly longer in patients who received HCQ compared to those who did not receive the treatment (17 [13–21] vs. 10 [4–13] days, p=0.023, Figure 1). The time to negativity test was not significantly different between patients with symptoms and without symptoms (14 [7–21] vs. 15 [4–21] days, respectively, p=1.00), patients who had pneumonia and those who had not (16 [12–21] vs. 13 [4–20] days, respectively, p=0.22), and patients who required oxygen therapy and those who did not (14 [9.2–21] vs. 14 [4.5–21] days, respectively, p=0.84).

**Figure 1:**
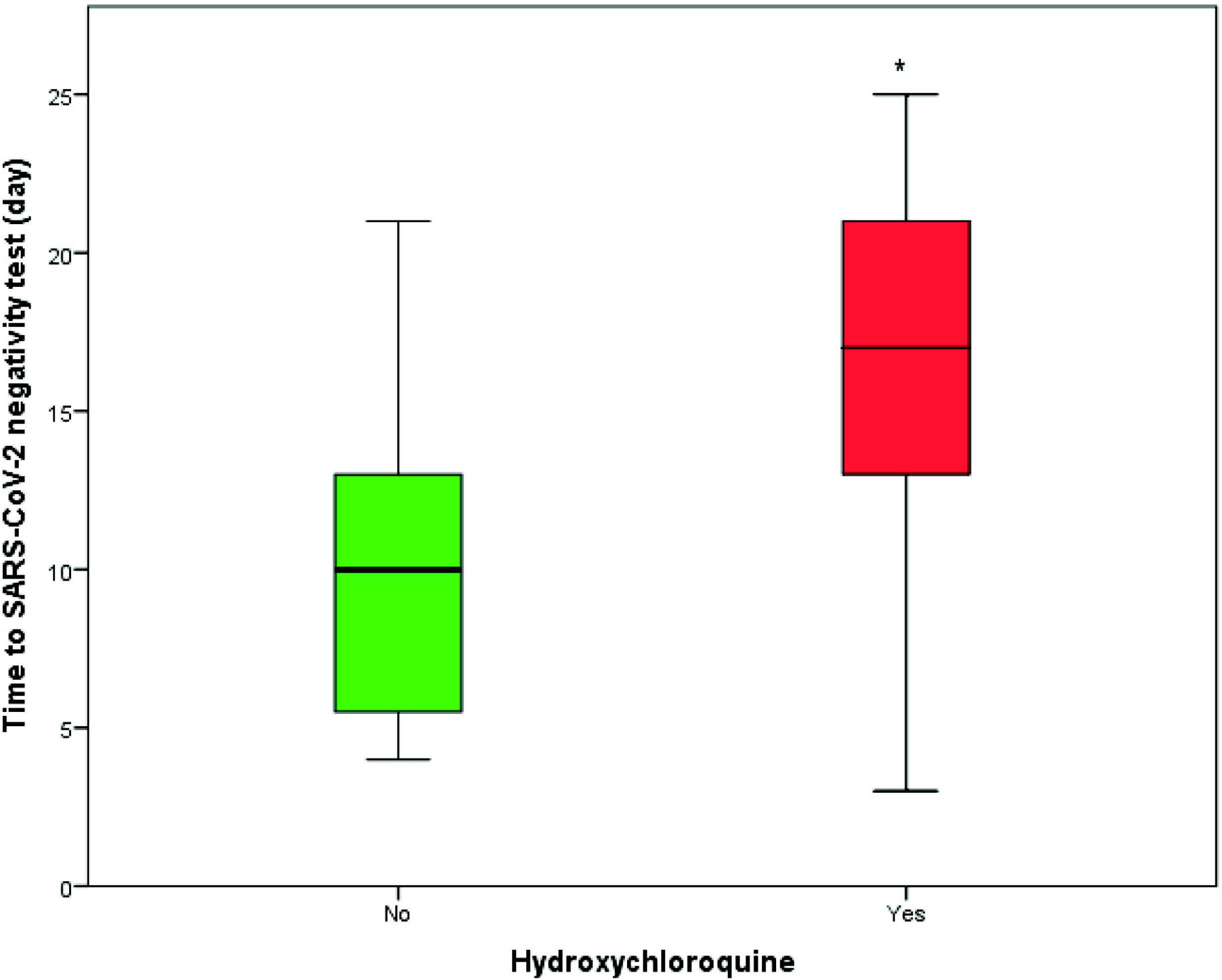
Time to SARS-CoV-2 negativity test is increased in hydroxychloroquine patients compared with patients who did not receive hydroxychloroquine. The median time to negativity test was 17 [13–21] days for hydroxychloroquine group and 10 [4–13] days for non-hydroxychloroquine group, ^*^ p=0.023.

Table 2 shows the results of the simple linear and multivariable regression analyses. No variables were significantly associated with the time to negativity test except for HCQ treatment in the simple regression analysis. After adjusting for these potential confounders: symptoms and pneumonia (Table 2) or symptoms and oxygen therapy (Table 1S), HCQ treatment was independently associated with a longer time to negativity test. For the reason of collinearity between oxygen therapy and pneumonia (p<0.001), these variables were not included together in the same multivariable model. On day 14, only 11 patients among the 23 patients treated with HCQ had their SARS-CoV-2 tests turned negative compared to 10 patients among the 11 patients who did not receive HCQ treatment (47.8% vs. 90.9%, respectively, p=0.016).

**Table 2.** Simple and multivariable linear regression analysis with time to negativity as a dependent variable

| Variables | Simple linear regression |  |  | Multivariable linear regression |  |  |
| --- | --- | --- | --- | --- | --- | --- |
|  | Beta coefficient | 95% CI | p-value | Beta coefficient | 95% CI | p-value |
| HCQ (reference: no) | 5.60 | 0.89-10.08 | **0.023** | 5.18 | 0.10-9.61 | **0.031** |
| Symptoms (reference: no) | 0.028 | -4.93-5.00 | 0.99 | 1.54 | -2.72-6.00 | 0.54 |
| Pneumonia (reference: no) | 3.36 | -1.90-8.22 | 0.18 | 2.92 | -1.73-7.74 | 0.24 |

HCQ, hydroxychloroquine; CI, confidence interval

### Effects of HCQ treatment on the time course of inflammatory markers

Table 3 shows that leucocytes counts, lymphocytes counts, lymphopenia rate, C-reactive protein, and ferritin did not significantly change between hospital admission and day seven or hospital discharge in the HCQ group nor the non-HCQ group.

**Table 3.**
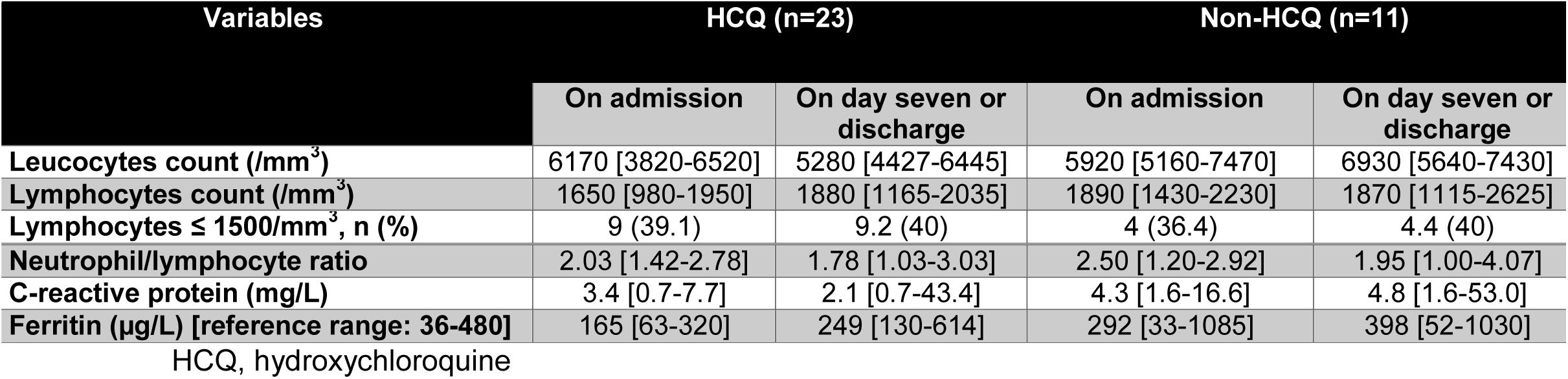
Time course of inflammatory variables between admission and day seven or hospital discharge in the HCQ and non-HCQ groups

## Discussion

The main findings of our study can be summarized as follows: (1) HCQ treatment was independently associated with a longer time to SARS-CoV-2 test negativity; (2) At day 14, virologic clearance was significantly higher in patients who did not receive HCQ; (3) HCQ treatment did not result in improvement of inflammatory markers or lymphopenia rate (Table 3).

HCQ has been widely used in the prevention and treatment of malaria and the treatment of chronic inflammatory diseases [15,16]. In-vitro studies have demonstrated that HCQ exhibits a non-specific antiviral activity and can block SARS-CoV-2 entry to cells through inhibiting the acidification of endosomes, which prevents membrane fusion and endocytosis of the viral envelop [6,7]. In a recent open-label nonrandomized study of 36 patients [8], Gautret et al. reported improved virologic clearance with HCQ compared to control patients receiving standard supportive care. Virologic clearance, measured by nasopharyngeal swabs, at day 6 was 57% (8/14) for patients who received HCQ monotherapy for 10 days compared to 12.5% (2/16) for patients who did not receive HCQ. In a recent study [9], the same authors, in a cohort of 80 confirmed COVID-19 patients with mild illness, observed that the combination of HCQ and azithromycin for 10 days resulted in reduced nasopharyngeal viral load (83% and 93% tested negative on days 7 and 8, respectively). Our findings stand in contrast with those reported by Gautret et al. [8,9] and cast doubt about the strong antiviral efficacy of HCQ. Indeed, we observed that HCQ was independently associated with a longer time for a positive nasopharyngeal swab to turn negative after adjustment for potential confounders (Table 2 and 1S), suggesting a slower viral clearance. Furthermore, a significantly higher percentage of our patients who did not receive HCQ tested negative on day 14 compared with those who received HCQ (90.9% vs. 47.8%, respectively). The studies reported by Gautret et al. [8,9] had major limitations. In the first study [8], 6 (23%) patients in the HCQ group were removed from the analysis due to early cessation of treatment resulting from critical illness (transfer to ICU) or intolerance of the drugs. Also, no safety or clinical outcome was reported. The second study [9] had no control arm. Our findings are partly in line with other studies that found no effects of HCQ on viral clearance [11,12,17]. In a prospective study of 30 COVID-19 patients [11], the authors randomized patients to HCQ (400 mg daily for 5 days) and standard of care or standard care alone. They found no significant difference in the rate of virologic clearance at day 7 between patients with or without HCQ treatment (86.7% vs. 93.3%, respectively), and no difference in clinical outcomes. Molina et al. [17], in patients who received HCQ for 10 days and azithromycin for 5 days, found that 80% (8/10) of them were still positive for SARS-CoV-2 in nasopharyngeal swabs 5 to 6 days after treatment initiation. In a recent multicenter, open-label randomized trial (preprint published) [12], 75 patients were assigned to HCQ (for 2 to 3 weeks) plus standard of care, and 75 patients were assigned to standard of care alone (control group). The authors found that the overall 28-day conversion rate (primary outcome) was not significantly different between the two groups (85.4% for HCQ group vs. 81.3% for control group). Also, the time to SARS-CoV-2 negativity test was not significantly different between HCQ and control groups (median 8 vs. 7 days, respectively).

Our study is the first to report a slower viral clearance with HCQ use in COVID-19 patients. Although there are no animal studies of chloroquine/HCQ in SARS-Cov-2 infection, data from other viral infections sometimes showed a deleterious effect on viral replication [18–20]. Chloroquine was shown to enhance alphavirus replication in various animal models [19,20] most probably because of the immune modulation and the anti-inflammatory effects of chloroquine in vivo [21]. Also, in a prophylactic study in a non-human primate model of chikungunya virus infection [18], chloroquine was shown to delay the cellular immune response, resulting in slower viral clearance. Furthermore, in a randomized, double-blind, placebo-controlled trial performed in 83 asymptomatic HIV patients [22], the use of HCQ compared with placebo resulted in a greater decline in CD4 cell count and increased viral load. Thus, it might be possible that the immunomodulatory effect of HCQ occasioned a slower clearance of the SARS-CoV-19 virus in our patients. However, this finding needs to be confirmed in further studies.

It has been reported that HCQ inhibits SARS-CoV-2 activity in-vitro with a half-maximal effective concentration (EC_50_) ranging from 4.5 µM to 17 µM [7], or 1507.5 µg/L to 5695 µg/L since the molar mass of HCQ is around 335 g/mole. Considering the blood volume of distribution of HCQ of 47257 Liters [16], 71240 mg of HCQ would be needed to be given (356 tablets of HCQ 200 mg!) to achieve an EC_50_ of 4.5 µM (1507.5 µg/L). Thus, it is unlikely that a standard dosing regimen of HCQ used in clinical practice would be able to inhibit viral activity in COVID-19 patients.

The use of HCQ did not result in the improvement of inflammatory parameters or the lymphopenia rate within seven days of admission. This might be explained by the low inflammatory reaction in our patients on admission suggestive of mild illness severity. Tang et al. [12] observed a significant decline in C-reactive protein levels in patients treated with HCQ. However, in their study, HCQ was given at much higher doses (1200 mg daily for 3 days, followed by 800 mg daily) than in our study. Even with such high dosing regimen, HCQ was not able to significantly increase the lymphocyte count [12].

HCQ use was well tolerated in our patients; we did not observe any side effects. This might be attributed to the low dosing regimen used in our study (400mg daily).

We acknowledge several limitations to our study, including small sample size and those inherent to retrospective designs. However, baseline patients’ characteristics and laboratory data were well balanced between HCQ and non-HCQ groups. Despite multivariable analysis and adjustment for potential confounders, we cannot rule out bias selection or residual confounding. We included patients with mild to moderate illness. Thus, our results cannot be applied to COVID-19 patients with severe disease.

## Conclusions

Despite a reported antiviral activity against SARS-CoV-2, we found that HCQ was associated with a slower viral clearance in COVID-19 patients with mild to moderate disease. Data from ongoing randomized clinical trials with HCQ should provide a definitive answer regarding the efficacy and safety of this treatment. Until then, the findings of our study suggest caution in using HCQ in hospitalized COVID-19 patients with mild to moderate illness.

**Author contributions:** Mallat had full access to all of the data in the study and takes responsibility for the integrity of the data and the accuracy of the data analysis.

*Concept and design:* Mallat, Hamed.

*Acquisition, analysis, or interpretation of data:* Mallat, Hamed, Bonilla, Balkis, Mooty, Mohamed, Malik, Nusair.

*Drafting of the manuscript:* Mallat

*Critical revision of the manuscript for important intellectual content:* Mallat, Hamed, Bonilla, Balkis, Mooty, Mohamed, Malik, Nusair.

*Statistical analysis:* Mallat.

**Conflict of Interest Disclosures:** None reported.

## Data Availability

Anonymized datasets can be made available on reasonable request after approval of our Ethics Committee.

---

**Table 1S.** Simple and multivariable linear regression analysis with time to negativity as a dependent variable

| Variables | Simple linear regression |  |  | Multivariable linear regression |  |  |
| --- | --- | --- | --- | --- | --- | --- |
|  | Beta coefficient | 95% CI | p-value | Beta coefficient | 95% CI | p-value |
| **HCQ (reference: no)** | 5.60 | 0.89-10.08 | **0.023** | 5.68 | 1.05-10.08 | **0.019** |
| **Symptoms (reference: no)** | 0.028 | -4.93-5.00 | 0.99 | 0.64 | -4.50-6.65 | 0.80 |
| **Oxygen therapy (reference: no)** | 0.75 | -4.57-6.37 | 0.76 | 0.86 | -3.38-5.85 | 0.68 |

HCQ, hydroxychloroquine; CI, confidence interval

## Notes

### Competing Interest Statement

The authors have declared no competing interest.

### Funding Statement

No external funding was received.

